# Identification of Respiratory Diseases using Peak Energy Analysis of Acoustic Cough

**DOI:** 10.1101/2024.05.29.24308077

**Authors:** Sujith Thomas Chandy, Balamugesh Thangakunam, Gowrisree Rudraraju, Narayana Rao Sripada, Jayanthy Govindaraj, Charishma Gottipulla, Baswaraj Mamidgi, Shubha Deepti Palreddy, Nikhil kumar Reddy Bhoge, Harsha Vardhan Reddy Narreddy, Prasanna Samuel P, Devasahayam Jesudas Christopher, Manmohan Jain, Venkat Yechuri

## Abstract

**Background and Objectives:** Cough is a common symptom of respiratory diseases and cough intensity analysis can act as a determinant for identifying a pathological condition in the lungs. Recent advancement on the analysis of the cough sound has suggested that it has the potential to be used as a non-invasive marker for screening respiratory conditions, such as Asthma, Chronic Pulmonary Obstructive Disease (COPD), Interstitial Lung Disease (ILD) and Bronchiectasis. Peak analysis of the energy envelope helps in quantifying the feature variation for these conditions.

This study highlights the variation of the peak energy features of cough sounds in different respiratory conditions.

**Methods and Materials:** For validation 1849 voluntary cough sounds were recorded through a mobile application from 1402 patients (both male and female) and made a respiratory sound database containing 851 normal, 510 Asthma, 80 Bronchiectasis, 252 COPD, and 156 ILD cough sounds. The cough sounds were labeled with corresponding pathologies from spirometry. From every subject three audio recordings were collected before taking a spirometry test. Peak analysis is performed on the features extracted from cough audio signals. Peak features are extracted using the function ^“^ scipy.signal.find_peaks^”^ from python^’^s Scipy library. The comparison of these features is done against the clinical diagnosis which the physician finally arrives at after going through the history, spirometry and radiology as per the standard of practice.

**Results:** The peak analysis in Asthma shows higher base distance and peak height than that of Normal because of prolonged expulsion and airways constriction. The base distance is observed high in Bronchiectasis, but peak height and prominence are less when compared with Normal due to loss of elasticity in the airways. Whereas in COPD the base distance and prominence are found to be less than Normal, Asthma and Bronchiectasis which is attributed to multiple narrowing of the glottis.

**Conclusion:** Peak analysis of cough provides inferences which can be used as descriptors to differentiate coughs related to respiratory diseases.

□ **What is already known on this topic** – Patients with respiratory diseases have cough as common symptom. Detailed assessments of cough help in measuring the transmission of respiratory infection, assisting in screening respiratory diseases.
□ **What this study adds** – Cough peak energy features act as indicators in identifying the type of respiratory disease just by recording voluntary cough sounds.
□ **How this study might affect research, practice or policy** – Peak energy features might contribute to the ongoing efforts to enhance diagnostic capabilities, assessments and management of respiratory health.

## Introduction

In recent years, the healthcare industry has seen many innovations in the form of cutting-edge technologies and cutting-edge treatment approaches. Developing predictive and representative health diagnosis systems is essential for continuous monitoring of symptoms to prevent diagnostic mistakes [1]. Recently, focus has shifted to data-driven processes and intelligent software, where tools based on computer-aided detection (CAD) and artificial intelligence (AI) are becoming increasingly popular [2]. While there is a proven track record of AI based algorithms in image-based diagnosis, voice-based diagnosis is getting more attention and making significant strides as sounds carry the signature of many diseases [3, 4].

Cough is a common symptom of several respiratory diseases. Any changes in the cough sound can reflect pathological changes in the lungs. In recent years, advancements in AI techniques have spurred numerous studies aiming to categorize respiratory diseases through the analysis of cough sound [5]. The audio features in cough signals usually contain information about predicament of the respiratory system in different pathological conditions [6]. The acoustic analysis of the cough sounds critically detects various respiratory maladies such as COVID-19 [7], chronic obstructive pulmonary diseases (COPD) [8], asthma [9–12], pertussis [13, 14], croup [15], bronchitis [16], pediatric pneumonia [17], lower and upper respiratory tract diseases [18]. Although numerous studies have illustrated that acoustic cough related features can be used to differentiate several respiratory diseases nevertheless, there are only a few correlations between acoustic characteristics and perception ratings. This study aims to quantify the feature variation of various respiratory conditions through the peak analysis of the energy envelope.

Peak energy analysis involves extracting relevant features from the cough sounds. Chatrzarrin et al., 2011 [19] proposed peaks of the energy envelope and spectral features of the cough sound to distinguish between wet and dry cough. This study emphasizes the energy associated with cough sound peaks as a primary feature for peak analysis. The peak energy can be quantified using techniques like root mean square (RMS) energy calculation. RMS is used to characterize the energy contained in the sound waves of a given cough segment.

This is substantiated by the study done by Ren et al., 2022 [20] which showed variations in the mean inter-peak distance in the root mean square energy (RMSE) between COVID – 19 negative and positive patients. Similarly, Ashby et al., 2022 [21] has proposed RMSE peak analysis as a cough audio segmentation framework for COVID detection. The RMSE was computed from Short-Term Energy (STE), which is the energy of a signal corresponding to the total magnitude of the signal. Further, the peak energy (peak height) calculated from RMS was an important parameter for investigating the relationship between cough sound and physiological measures of cough strength [22]. This shows the weightage of the acoustic features both in conventional and machine learning models. A study carried out by Chung et al., 2021 [23] emphasized the role of loudness and energy ratio as the objective characteristic indicator in the acoustic analysis of cough sounds. In the study, the newly identified characteristics of the cough sound like energy ratio that effectively reflected the acoustic characteristics of pneumonia cough. The accuracy of the ML system to predict disease by combining the time-domain feature like peak energy with the frequency domain feature is well established in a study conducted by Ismail et al., 2022. The importance of peak energy analysis is well articulated in the study, which implemented it as a part of mixed-domain feature vector for the quick identification or early screening of COVID-19 [24]. Apart from the identification of pathological cough, the peak energy analysis along with the other time domain and frequency domain features was used to clearly distinguish between coughs and other noises in a sleep environment [25–30].

Based on the features extracted in our previous work, this study would provide insights on variation of the peak energy features on the RMSE envelope of cough sounds. By extracting and scrutinizing the peak properties of energy contours like peak height, peak prominence, base distance (distance between left and right base of a peak), the study aims to unveil intricate details pertaining to cough patterns associated with diverse respiratory diseases. The study also aims to create a strong framework for measuring and distinguishing cough patterns by combining these peak energy features and examining their correlation with various pathological conditions.

## Materials and methods

### 1. Cough dataset

Patients who visited an outpatient clinic at Christian Medical College (CMC) Vellore, Tamil Nadu, India, were enrolled in the clinical study (CTRI/2021/04/032742). The objective of the study is to validate the Swaasa.AI platform for screening and diagnosis of respiratory diseases based on audiometric evaluation of voluntary coughs sounds using machine learning algorithm. To perform an assessment using Swaasa® AI platform, patients’ demographic details and vital signs were collected, followed by interviews using the respiratory questionnaire to gather information on respiratory symptoms and lung health. Trained healthcare personnel recorded cough sounds using smartphones, ensuring standardized procedures such as maintaining distance, angle, and duration. The recording proceeded in the actual clinical field (outpatient clinic). The collected cough sound recordings “wav files” were then subjected to noise filtering techniques, tailored to the device type. All sounds were carefully checked and validated by pulmonologists.

All patients gave written informed consent, and we obtained human research ethics committee approval of Christian Medical college Institutional Review Board. All methods were performed in accordance with the relevant guidelines and regulations. We recorded 1849 cough sounds from 1402 patients (both male and female) and made a respiratory sound database containing 851 normal, 510 Asthma, 80 Bronchiectasis, 252 COPD, and 156 ILD cough sounds. Characteristics of the cough sound database are detailed in table 1.

**Table 1:** Characteristics of cough sound database.

| 1402 subjects (cough sounds: n=1849) | Characteristics |  |
| --- | --- | --- |
| Age |  |  |
|  | 18 – 47 years | 1005 (54.4) |
|  | 48 – 63 years | 575 (31.1) |
|  | 64 – 89 years | 269 (14.5) |
| Sex |  |  |
|  | Male | 1145 (61.9) |
|  | Female | 704 (38.1) |
| Disease |  |  |
|  | Normal | 851 (30.1) |
|  | Asthma | 510 (18.0) |
|  | Bronchiectasis | 80 (2.8) |
|  | COPD | 252 (9.0) |
|  | ILD | 156 (5.5) |

We arrived at this number by using a statistical method, based on the assumption that the device could present a sensitivity and specificity of 80% with an error of 5% for 95% CI in the detection of respiratory conditions, and a prevalence of the same of 25%, a sample size of 1500 sounds was found to be adequate for validation. However, to ensure rigor and improve the accuracy of the estimates, a larger sample size of 1849 was estimated for a sensitivity/specificity of 90%.

To ensure consistency all over the dataset, preprocessing of three major sound properties (Audio Channels, Sample Rate) was done. The audio channels of the cough samples were integrated into mono channels and the sample rates were modified to the default sample rate of 16000 kHz.

### 2. Acoustic analysis and Peak analysis of the Cough sound

#### 2.1. Acoustic Analysis

Acoustic analysis provides an objective means of evaluating the sound characteristics of coughs. Investigations into waveforms of voluntary cough sounds known as, ‘Tussi phonograms’, indicate the potential diagnostic utility. However, comprehensive validation studies for this purpose are lacking [23]. Exploring the acoustic properties of voluntary cough sounds have revealed notable distinctions among coughs associated with various diseases [24]. Analyzing the waveforms which capture time and frequency content, can help identify characteristics in cough sounds that are indicative of the presence of mucus in the airways [25, 26].

In our study, we have analyzed Root Mean Square Energy (RMSE) envelopes extracted from cough sounds by quantifying the peak features. RMSE in an audio signal represents the average power or intensity of a signal over time. It’s a statistical measure that considers all values in a signal, not just the peaks, to provide a more accurate representation of the signal’s energy. It provides a measure of the perceived loudness of an audio signal, considering the entire time range. It can help in analyzing the intensity and variations in loudness over time.

#### 2.2. Audio cough recording

Audio cough recordings are captured using a smartphone which has the following specifications.

Audio frequency range 20Hz-20kHz,

Sensitivity: −26 dB to +/−2dB

Maximum sound pressure level: 98dB - 120 dB SPL

Dynamic range: 60dB.

Here is the cough audio recording of an ILD male participant with a high severity restriction pattern, where the duration of the record was about 10 seconds (Fig 1a). It was found that the 10 second (s) cough sound had 8 cough sequences which were labeled from S1 to S8 respectively. The extrapolation of the cough sequence is shown in fig 2b, where the sequence 2 (S2) was found to have three coughs – C1, C2 and C3. In the above record for 8 sequences there are 27 coughs (this is a scenario of high cough count). The various features are extracted and analyzed from the cough sounds.

**Figure 1a, 1b & 1c.**
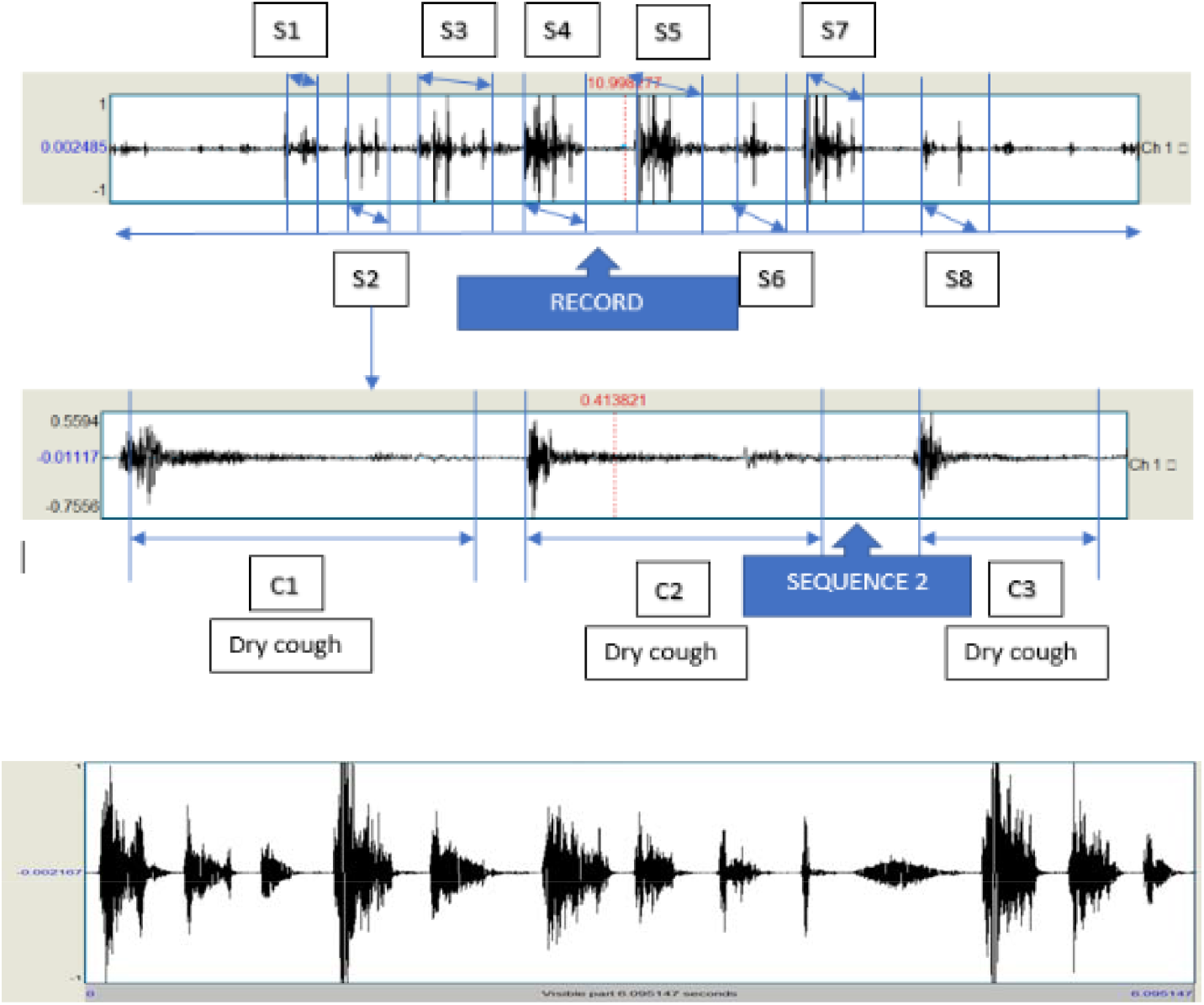
Audio cough recording of 10 seconds. S1-S8 – Sequences. C1-C3 – Cough sounds.

**Figure 2:**
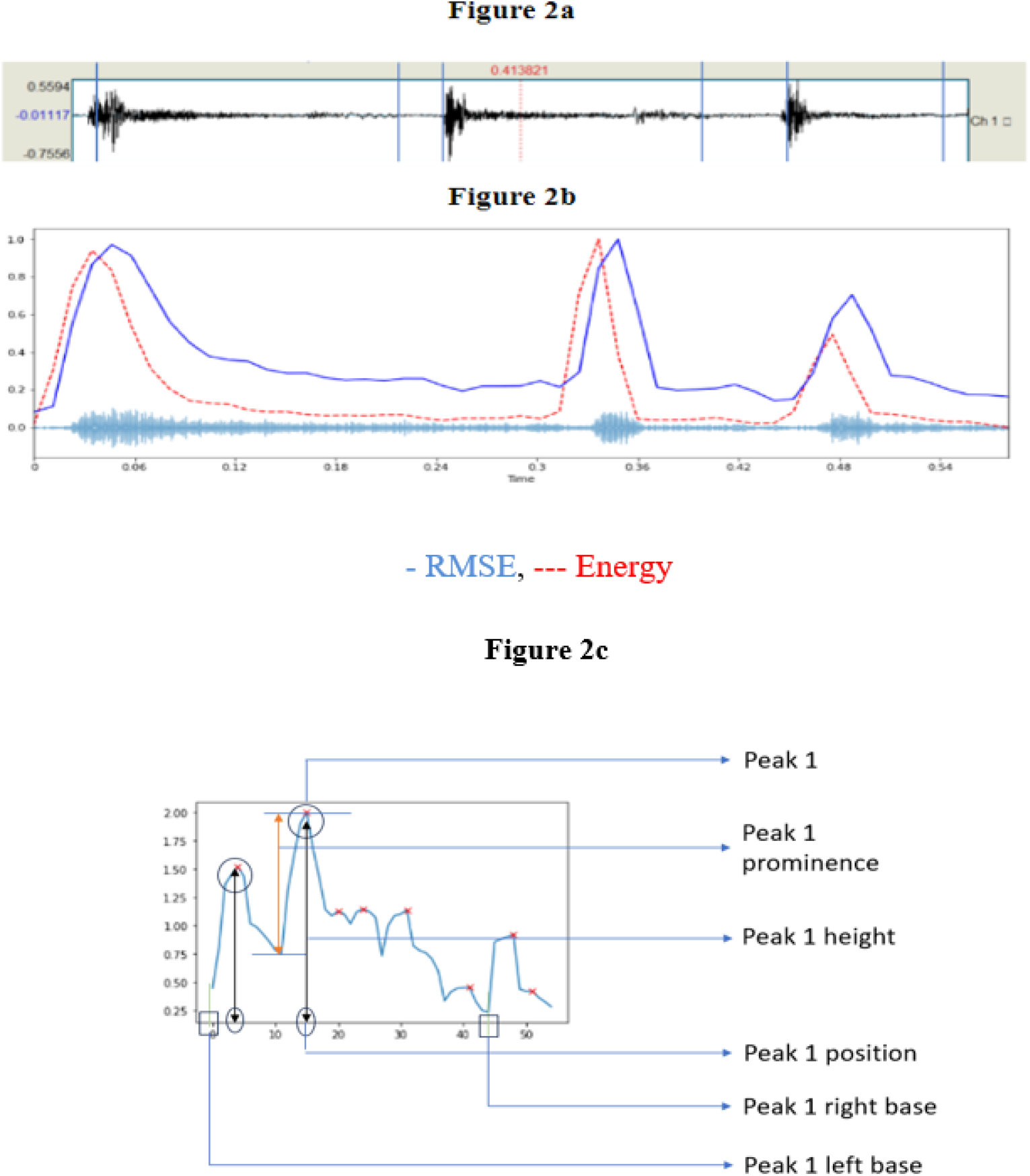
**2a**- Audio cough sequence; 2**b**- Acoustic cough feature - Root Mean Square Energy (RMSE) plot; **2c**- Various peak features derived from the RMSE plot

For peak analysis we have merged all the sequences extracted from the audio cough recording and considered the first six seconds from it.

#### 2.3. Peak Analysis

Peak analysis is performed on the features extracted from cough audio signals. Peak features are extracted using the function “scipy.signal.find_peaks” from python’s Scipy library. The peak features are obtained from peak properties, and they are -

- Peak height - Height of the peak (length).
- Peak prominence - The prominence of a peak measures how much a peak stands out from the surrounding baseline of the signal and is defined as the vertical distance between the peak and its lowest contour line.
- Peak left base - The peak’s base to the left.
- Peak right base - The peak’s base to the right.

Features such as Root Mean Square Energy (RMSE), Zero-crossing rate (ZCR), and Spectral centroid were extracted using signal standard processing techniques in both time and frequency domain. Peak analysis is performed on the envelopes of these features.

#### 2.4. RMSE peak analysis

Root Mean Square Energy (RMSE) is a time domain feature. Peak analysis is performed on the RMSE envelope. A time-domain graph shows how a signal changes with time, whereas a frequency domain graph shows how much of the signal lies within each given frequency band over a range of frequencies.

Figure 2a shows the actual signal and Figure 2b shows the RMSE envelope which is based on all samples in a frame (y-axis in volts and x-axis in frame numbers). It acts as an indicator of the energy, amplitude of the sound.

The prominence, height and the base distance of the peak was measured in the RMSE peak analysis for normal cough and different pathological conditions. Figure 2c shows the representation of the different features of the RMSE plot for a typical cough sound. It is observed that the base distance of the peak is measured in terms of right base and left base.

#### 2.5. Classifiers

We have built two classifiers, one with peak features included and another one without peak features. The difference in the performance of the classifiers has shown the effectiveness of peak features in identifying the respiratory disease. In this study, we reapply this well-proven feature set in combination with the peak energy features to investigate the basic feasibility of detecting respiratory diseases from cough sounds.

Cough sounds, have very complicated structures with noise, and positional dependency in time. In the sound analysis, particularly from a mathematical point of view, its 2-D spectral-domain has more information rather than one dimensional time-series. Moreover, the deep learning structure gives an automatic extraction overcoming the difficulties on complicated data, especially image data. For this reason, we adopted CNN, which is a powerful method in image classification.

Classifier 1 - The Convolutional Neural Network model was built based on transfer learning using Resnet-34 with imagenet for training on spectrograms. Whereas the Feed Forward Artificial Neural Network (FFANN) was utilized to process the tabular data (features extracted from 1-D time series). Tabular data includes time domain features like energy, zero crossing rates and frequency domain features like spectral centroid, spectral roll-off, spectral bandwidth, dominant frequency, spectral skewness, spectral kurtosis, spectral crest, spectral spread and spectral entropy. The last fully connected layers of both models were removed, and a new fully connected layer (merged layer) was added to predict the final output (type of respiratory disease) as shown in figure 3. This merging approach of the last layers of the two models was named the combined logic.

**Figure 3.**
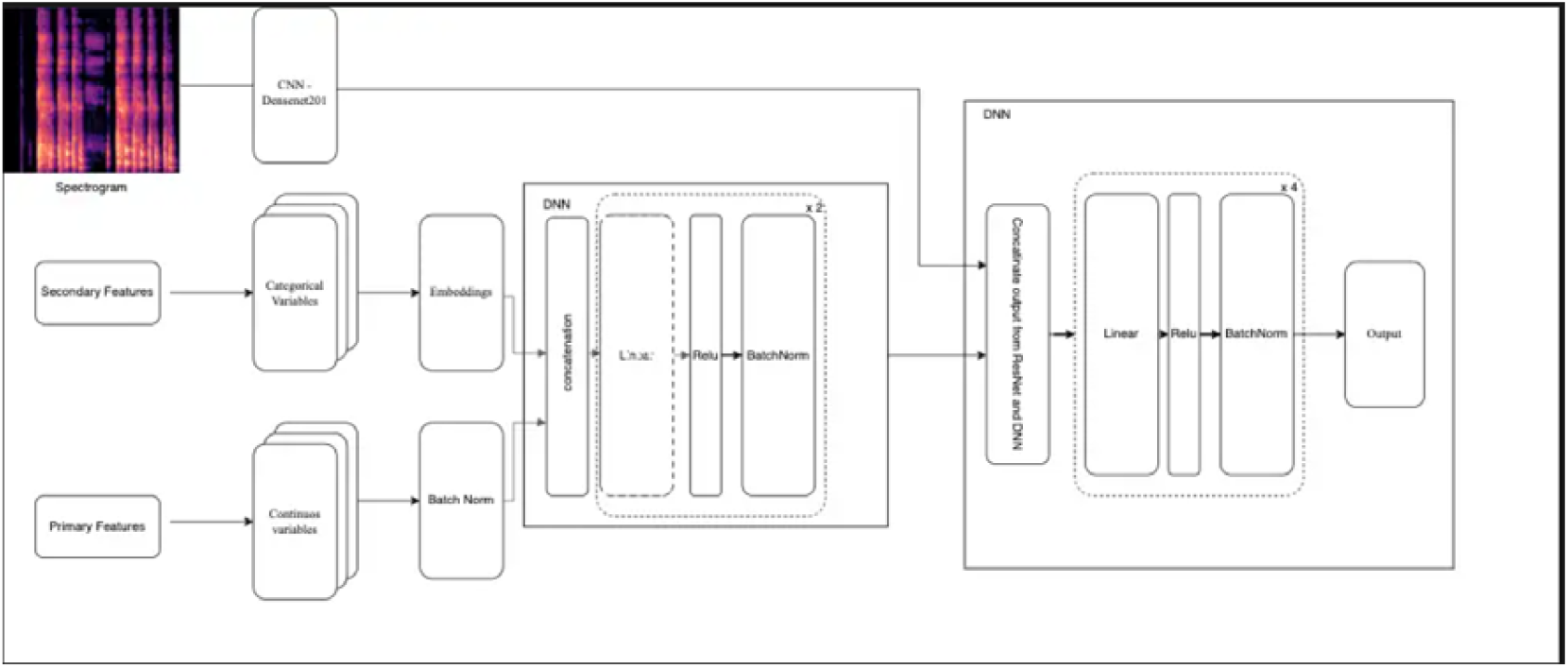
Architecture diagram of the machine learning model.

Classifier 2 – This classifier is the same as classifier 1 but peak energy features are added to the tabular data.

### 3. Results

#### 3.1 Peak analysis of cough sounds of different respiratory diseases

In female participants the mean peak height of the COPD cough was found to be lower (the average mean value of the peak feature in decibels relative to full scale (dBFS) is lower when compared) than the normal cough recorded for 6s. This decrease in the peak heights of the COPD cough may be attributed to the excess mucus and inflammation in the airways, destruction in the airways, resulting in less forceful expulsions of air during coughing. It was also observed that the severe condition of COPD exhibited lesser peak heights when compared to the medium and low severe conditions. This connotes the stage of advanced COPD where the respiratory function is significantly compromised.

For male participants, the peak heights of the COPD and the normal cough sound recorded for 6s was observed to have a similar pattern to that of the female subjects. Another interesting finding was that the COPD cough of the male subjects displayed more data points for lesser peak heights when compared to the COPD cough of the female subjects.

The table 2 below illustrates the comparison of peak height and base distance of coughs in different respiratory diseases with the help of box plots.

**Table 2:**
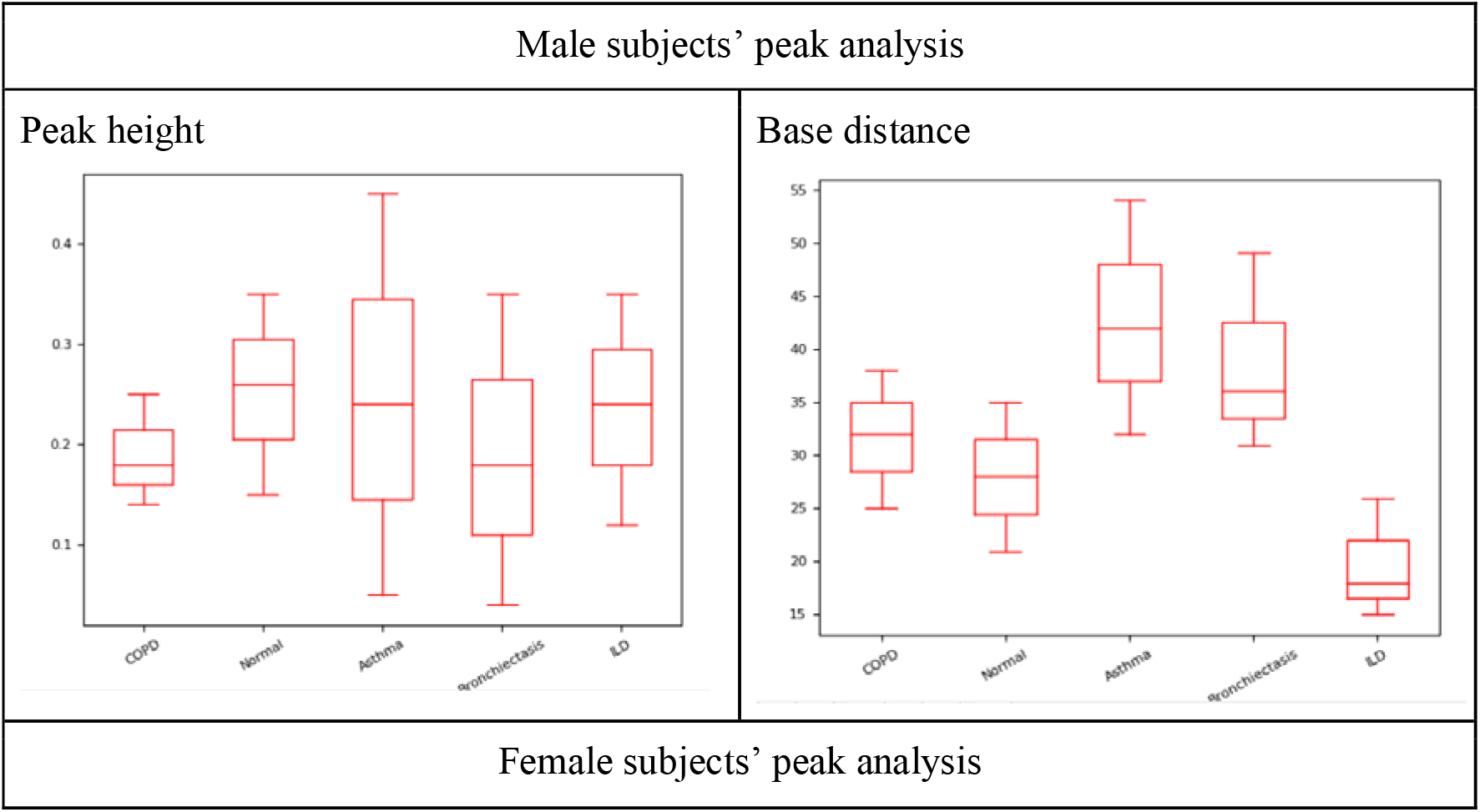
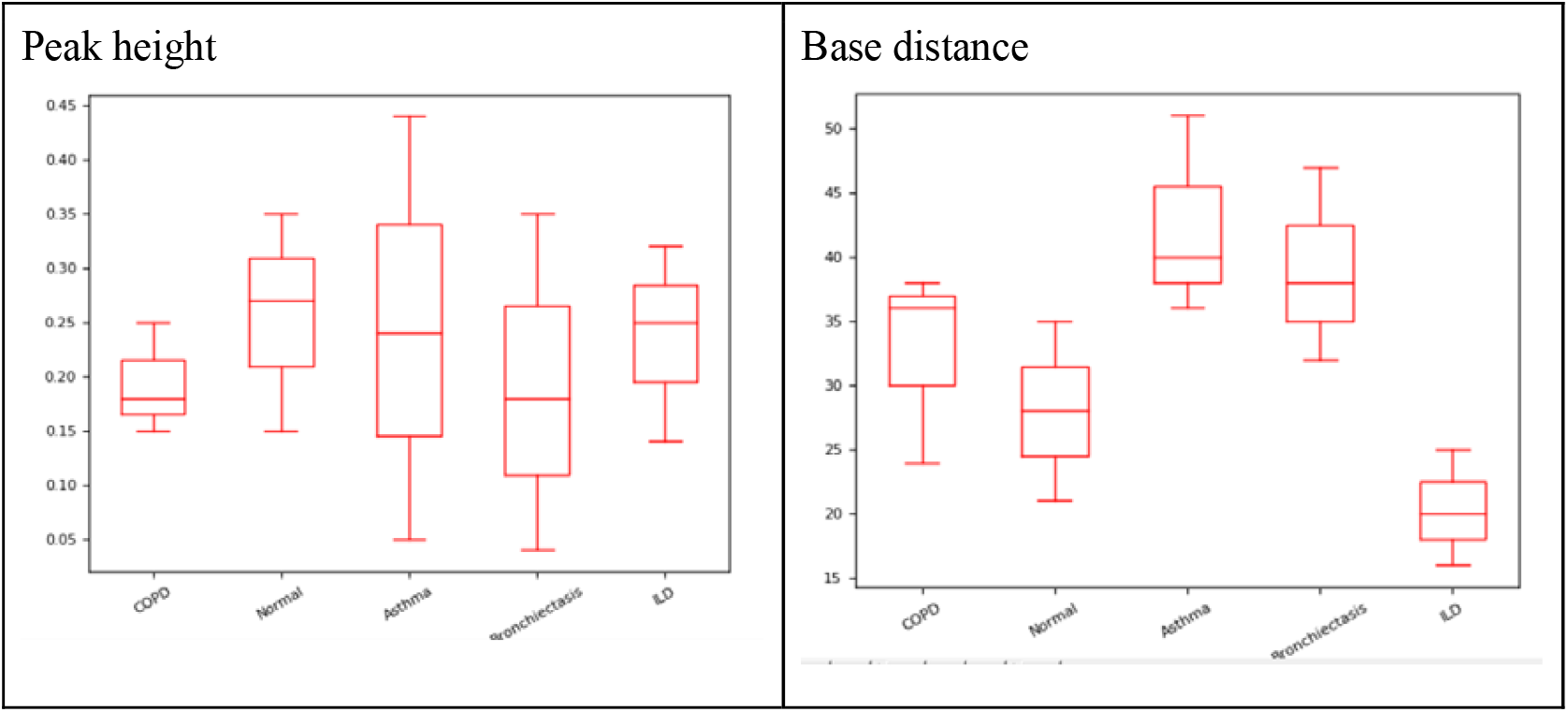
Peak analysis of female and male participants.

#### 3.2 RMSE base distance analysis

Base distance is the difference between right and left bases. The base distance gives the duration of the cough sequence.

In Table 3 we have captured base distances of different respiratory diseases. In Asthma due to airways constriction cough is produced with high energy and base distance (right base – left base) is very high because of prolonged expulsion.

**Table 3:**
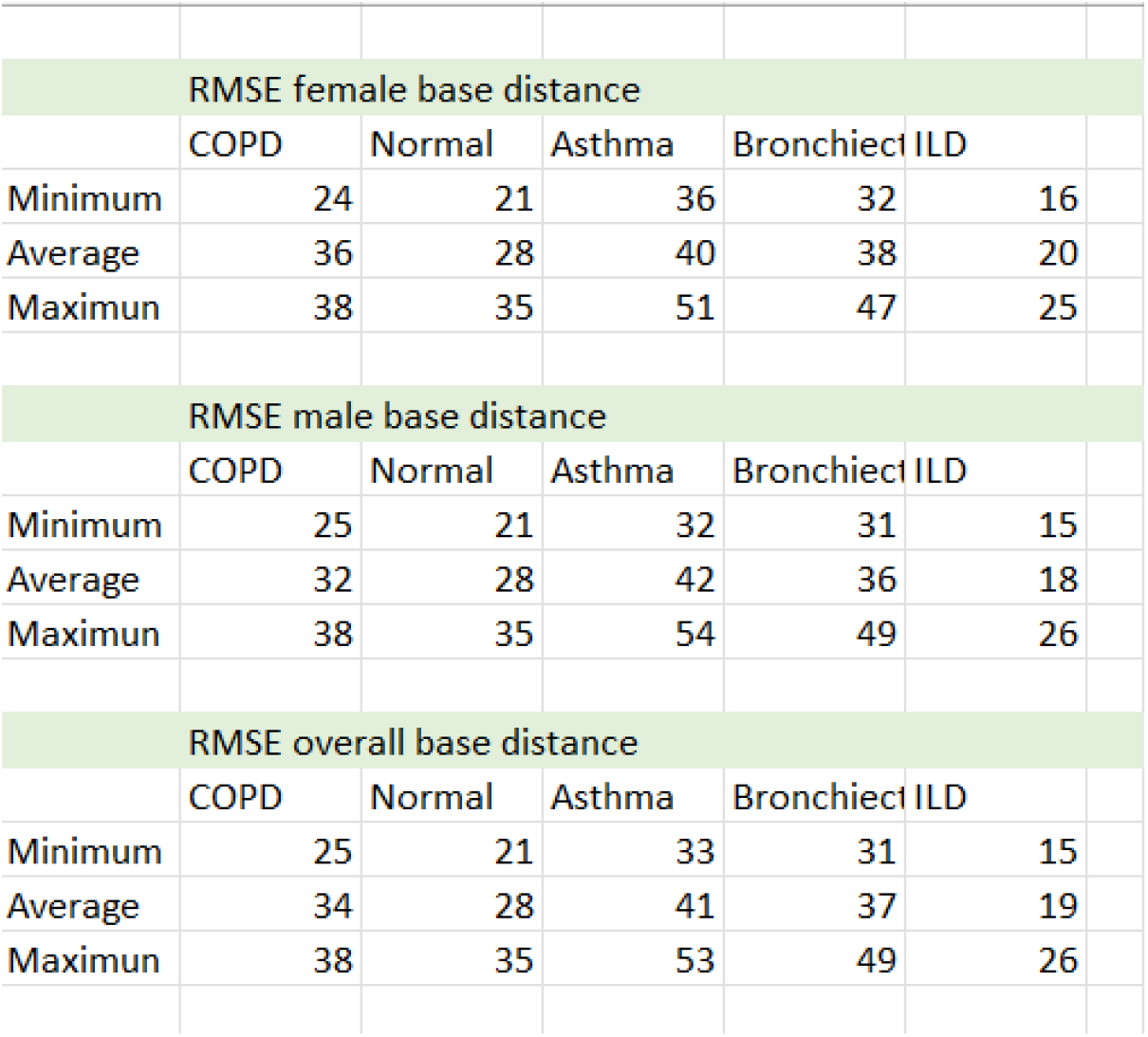
Base distance of female and male subjects in different diseases.

| RMSE female base distance |  |  |  |  |  |
| --- | --- | --- | --- | --- | --- |
|  | COPD | Normal | Asthma | Bronchiect | ILD |
| Minimum | 24 | 21 | 36 | 32 | 16 |
| Average | 36 | 28 | 40 | 38 | 20 |
| Maximun | 38 | 35 | 51 | 47 | 25 |
| RMSE male base distance |  |  |  |  |  |
|  | COPD | Normal | Asthma | Bronchiect | ILD |
| Minimum | 25 | 21 | 32 | 31 | 15 |
| Average | 32 | 28 | 42 | 36 | 18 |
| Maximun | 38 | 35 | 54 | 49 | 26 |
| RMSE overall base distance |  |  |  |  |  |
|  | COPD | Normal | Asthma | Bronchiect | ILD |
| Minimum | 25 | 21 | 33 | 31 | 15 |
| Average | 34 | 28 | 41 | 37 | 19 |
| Maximun | 38 | 35 | 53 | 49 | 26 |

COPD is a condition of inflammation/obstruction of airways; destruction of lung tissue/alveoli. Cough is prolonged and emerges as a reaction to clear the fluid (mucus), base distance is low due to dynamic narrowing of the glottis.

Bronchiectasis is a condition where inflammation permanently destroys the bronchial walls causing them to widen. The abnormally dilated bronchi filled with excess mucus, which can trigger persistent coughing and make the lungs more vulnerable to infection. Bronchiectasis also has high base distance but less when compared with asthma.

Normal **c**ough emerges as a defense mechanism; as the site of the irritant is mostly in large airways, pressure built inside is high, energy of the cough is also high, base distance is less.

#### 3.2 Comparison of peak analysis in different respiratory diseases

Cough peak energy features of different respiratory diseases are compared in table 4 below. Asthma cough is dry with high energy, Peak 1 height is high but less than normal. COPD cough has excess mucus; and has following conditions: inflammation in airways; destruction/enlargement of airways; destruction of lung tissue/air sacs. Hence, cough is prolonged and emerges as a reaction to clear the fluid (mucus). Due to presence of liquid - Prominence is very less, peak 1 and peak 2 heights are very close. Bronchiectasis can occur if the inflammation permanently destroys the elastic-like tissue and muscles surrounding the bronchi (airways), causing them to widen. The abnormal bronchi then becomes filled with excess mucus, which can trigger persistent coughing and make the lungs more vulnerable to infection. Peak height and prominence are observed to be less. Normal cough emerges as a defence mechanism; as the site of the irritant is mostly in large airways, pressure built inside is high, Energy of the cough is also high, duration of cough is less i.e distance between left base and right base of peak 1 is less, peak 2 height is also more prominence is also high.

**Table 4:**
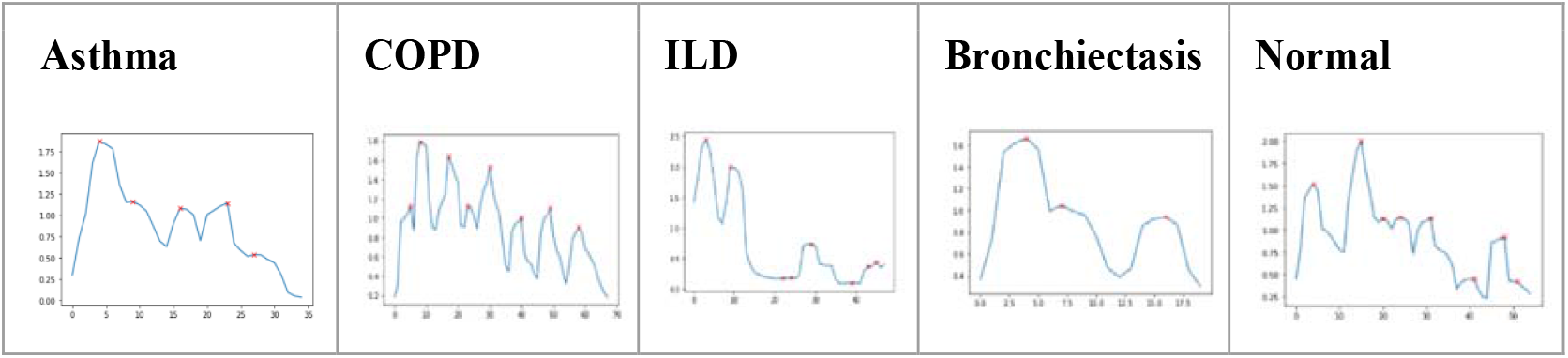
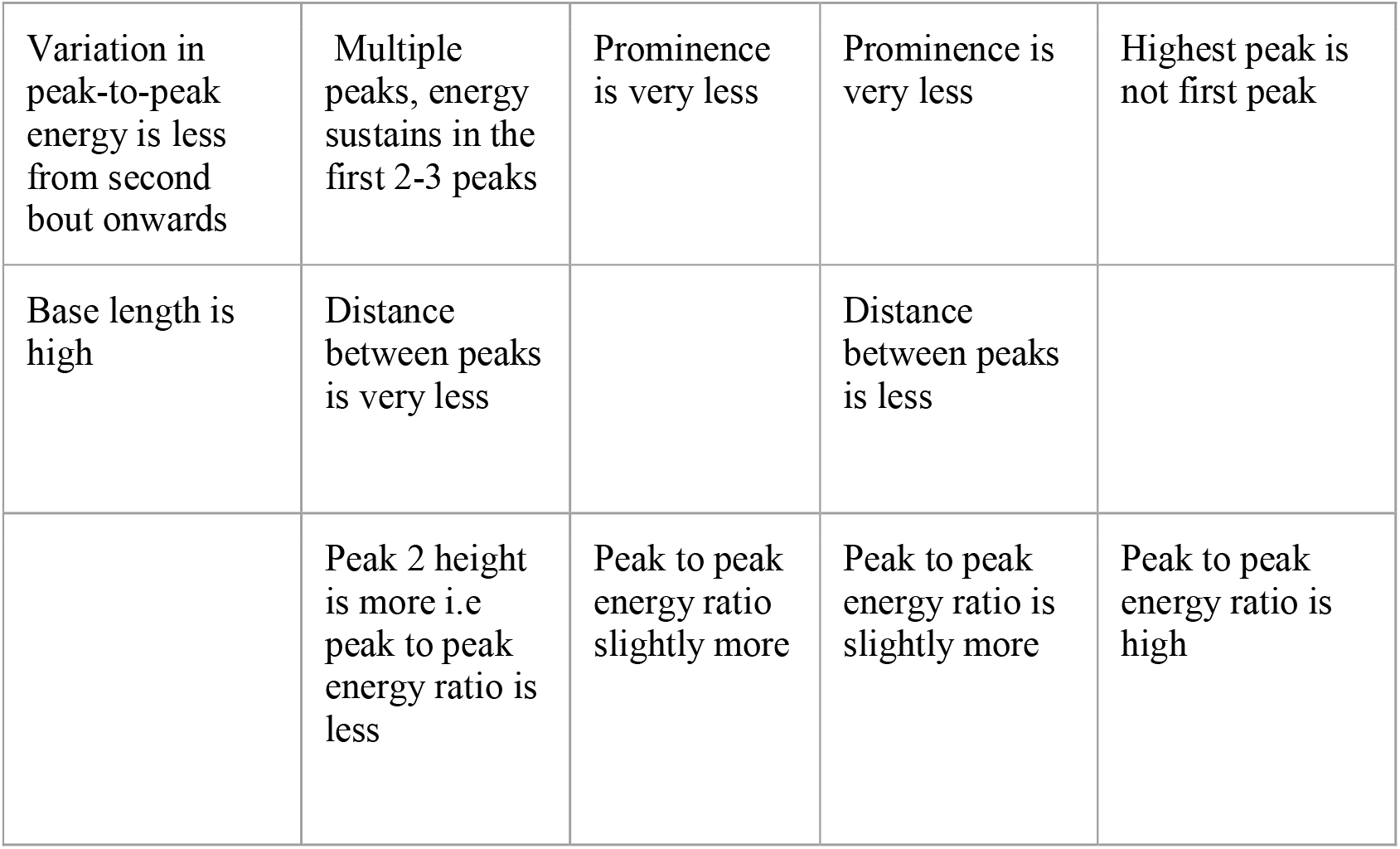
Comparing peak energy features of cough sounds in different respiratory diseases.

#### 3.3 Comparison of classifiers with and without peak energy features

The Disease model classifier is focused on detecting various respiratory conditions. Swaasa outputs diseases as “Asthma,” “COPD,” “ILD,” “Bronchiectasis,” and “Normal”. From the pool of 1917 subjects, 515 were chosen for derivation, and this dataset was integrated into the existing training dataset. The remaining 1402 subjects were employed for validating the performance of the disease models. It is essential to note that this segregation of data aligns with the specific input requirements necessary for training each model. Table 5 displays the confusion matrix of the classifier with peak energy features. Table 6a and 6b illustrates the performance metrics of the classifier with peak energies and without peak energies respectively.

**Table 5:** Confusion matrix of the classifier with peak features.

|  | Asthma | Bronchiectasis | COPD | ILD | Normal |
| --- | --- | --- | --- | --- | --- |
| Asthma | 348 | 18 | 100 | 19 | 25 |
| Bronchiectasis | 10 | 42 | 12 | 8 | 8 |
| COPD | 28 | 35 | 154 | 23 | 12 |
| ILD | 16 | 24 | 28 | 80 | 8 |
| Normal | 98 | 13 | 16 | 21 | 703 |

**Table 6a:** Performance of the classifier with peak energy features.

| Disease diagnosis type | Accuracy (%) | Sensitivity (%) | Specificity (%) |
| --- | --- | --- | --- |
| Normal | 89.1 | 82.6 | 94.7 |
| Asthma | 83.0 | 68.2 | 88.6 |
| Bronchiectasis | 93.1 | 52.5 | 94.9 |
| COPD | 86.3 | 61.1 | 90.2 |
| ILD | 92.1 | 51.3 | 94.8 |

**Table 6b:**
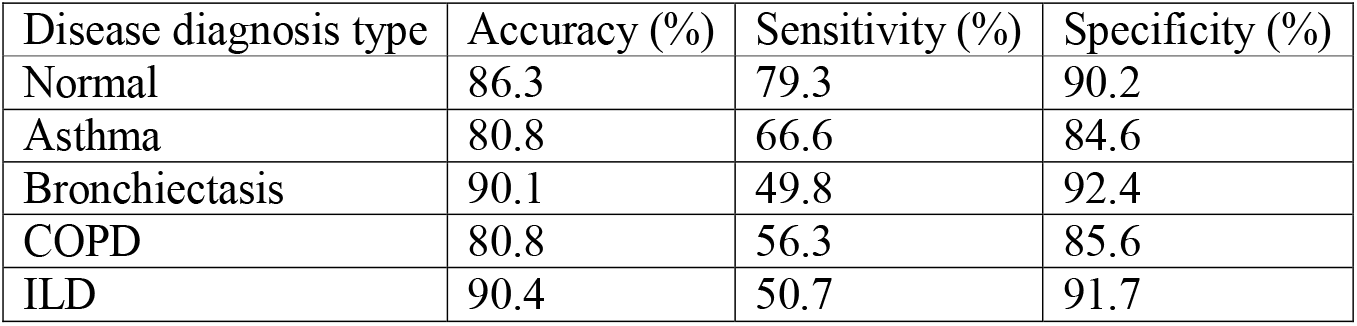
Performance of the classifier without peak energy features.

| Disease diagnosis type | Accuracy (%) | Sensitivity (%) | Specificity (%) |
| --- | --- | --- | --- |
| Normal | 86.3 | 79.3 | 90.2 |
| Asthma | 80.8 | 66.6 | 84.6 |
| Bronchiectasis | 90.1 | 49.8 | 92.4 |
| COPD | 80.8 | 56.3 | 85.6 |
| ILD | 90.4 | 50.7 | 91.7 |

The results of the classifiers were compared with diagnosis based on clinical diagnosis provided by the pulmonologist. The comprehensive evaluation of the model performance on the test set includes accuracy, sensitivity, specificity.

When compared to the existing classical classifier, peak energy features classifier gave better results in identifying respiratory diseases from voluntary cough audio recordings. Swaasa^®^ disease classifier with peak energy features commends a higher level of precision in respiratory diseases.

When compared with Asthma, ILD, and Bronchiectasis, COPD disease identification improved significantly (6%) with the addition of peak features.

## Discussion

The findings underscore the potential of RMSE peak analysis in discerning the normal and pathological conditions and distinguishing the severity of respiratory conditions. Analyzing both the male and female subjects in detail contributes to a holistic understanding of the variation of acoustic features across diverse demographics and respiratory conditions. Moreover, the exploration of peak height, peak prominence, and base distance as distinctive parameters for characterizing cough patterns provides nuanced understanding of the acoustic signatures associated with various respiratory conditions.

Additionally, the combination of spectrogram analysis and RMSE peak analysis also offers a multi-faceted approach to characterizing cough sounds. The correlation of the peak energy features with respiratory conditions may open avenues for more precise diagnostic tools and monitoring systems.

Thus, the findings of the study presented herein might contribute to the ongoing efforts to enhance diagnostic capabilities, fostering a new phase in the assessment and management of respiratory health. Our future studies might aim in the addition of peak analysis to ML models to increase accuracy in cough detection and thereby to explore the correlation between acoustic features and clinical parameters, enhancing the applicability of our findings in healthcare settings.

## 4. Limitations

One notable limitation of the study is the impediment encountered during peak analysis, primarily attributed to the presence of noise in the data, that might introduce complexity in accurately isolating and characterizing peak energy features.

## 5. Conclusion

In conclusion, our comprehensive analysis of cough sounds through spectrogram and RMSE peak analysis provides a novel foundation for the development of diagnostic tools for respiratory conditions, particularly COPD. The study also contributes valuable insights into the peak energy analysis of the acoustic characteristics of cough sounds, showcasing the potential for acoustic analysis as a non-invasive and informative tool in the assessment of respiratory health and improving clinical diagnostics.

## Data availability statement

Due to the nature of this research, participants of this study did not agree for their data to be shared publicly. However, the detailed analysis can be shared by the author “NRS” upon reasonable request.

## Ethical clearance

The study was registered under Clinical Trials Registry-India (CTRI/2021/04/032742) and was begun after getting the approval from the CMC-IRB (Institutional Review Board).

## Author Contributions

STC and BT defined study protocols, including study design and methodology. NRS conceptualized the idea of using cough sounds for screening and diagnosing COVID-19. GR performed literature review and data analysis. BM, HVR, SDP and NKRB were involved in device development. VY and MJ created value propositions for the device. SS assisted in executing the project at Christian Medical College by providing all the resources and extending research capabilities. CG and GR performed data analysis, sample size estimation and result analysis. KLPK, SS, NJ, VSP, ST and SV provided subject matter expertise. GR and JG wrote the manuscript. All the authors provided intellectual inputs and helped in preparing the manuscript.

## Competing interest statement

The authors have no conflicts of interest to declare. All co-authors have seen and agree with the contents of the manuscript and there is no financial interest in reporting. We certify that the submission is original work and is not under review at any other publication.

## Notes

### Competing Interest Statement

The authors have declared no competing interest.

### Clinical Trial

CTRI/2021/04/032742

### Funding Statement

This study is supported by the UK Government (British High Commission, New Delhi). This is a commissioned research report on commercial terms between C-CAMP and the UK Government (British High Commission, New Delhi).

### Author Declarations

IRB of Christian Medical College gave ethical approval for this work.

### Summary of Updates

Added and modified tables. Elaborated the methods in the tables.

